# Sex differences and Psychological Stress: Responses to the COVID-19 epidemic in China

**DOI:** 10.1101/2020.04.29.20084061

**Authors:** Kangxing Song, Rui Xu, Terry D. Stratton, Voyko Kavcic, Dan Luo, Fengsu Hou, Fengying Bi, Rong Jiao, Shiyan Yan, Yang Jiang

**Author notes:** Corresponding author: Yan Shiyan, School of Acupuncture-Moxibustion and Tuina, Beijing University of Chinese Medicine, Beijing 100029, China, 13521436209. Kangxing Song and Rui Xu have the equal contribution on the study.

## Abstract

About 83000 COVID-19 patients were confirmed in China up to May 2020. The effects of this public health crisis - and the varied efforts to contains its spread - have altered individuals’ “normal” daily functioning. This impact on social, psychological, and emotional well-being remain relatively unexplored, especially the ways in which Chinese men and women experience and respond to potential behavioral-related stressors. A cross-sectional study was conducted in late February 2020. Demographic characteristics and residential living conditions were measured along with psychological stress and behavior responses to the COVID-19 epidemic. 3088 questionnaires were received: 1749 females (56.6%) and 1339 males (43.4%). The mean level of stress, as measured by a visual analog scale, was 3.4 (SD=2.4) - but differed significantly by sex. Besides sex, factors positively associated with stress included: age (≤45 years), employment (unsteady income, unemployed), risk infection population (exposed to COVID-19, completed medical observation), difficulties encountered (diseases, work/study, financial, mental), behaviors(higher desire for COVID-19 knowledge, more time spent on the COVID-19). “Protective” factors included frequently contact with colleagues, calmness, and psychological resilience. Males and females also differed significantly in adapting to current living/working status, coping with heating, and psychological support service needs. Among Chinese, self-reported stress related to the COVID-19 epidemic were significantly related to sex, age, employment, resilience and coping styles. Future responses to such public health threats may wish to provide sex- and/or age-appropriate supports for psychological health and emotional well-being to those at greatest risk of experiencing stress.

## Introduction

Novel coronavirus disease 2019 (COVID-19) was now classified by the World Health Organization (WHO) as a full-blown pandemic. In this COVID-19 epidemic, there were about 83000 patients were confirmed in China and a series of strictly control measurements altered individuals’ “normal” daily functioning were adopted[1]. In the public eye, a lack of effective therapy, overlong incubation, asymptomatic infection, and talk of “post-recovery” reinfections exacerbated an already tense and uncertain situation[2-4], Meanwhile, efforts to contain the spread - such as city lockdowns, home quarantines, and suspension of public and private services-resulted in behavioral changes impacting individuals’ social, psychological, and economic well-being[5].

As “ground zero” for the ensuing epidemic, the ongoing COVID-19 threat is thought to have exacted a particularly dramatic psychological toll on the Chinese population. Indeed, a Hong Kong survey conducted during the early phases of the epidemic indicated that 97% of respondents were worried about COVID-19 - and 99% were alert to the disease progression, associated anxiety levels, and high perceived susceptibility and severity[6]. Similarly, medical workers reported increased stress, anxiety, and depressive symptoms associated with the emergence, spread, and potential lethality of the virus[7,8].

The potential impact of stress on anxiety, depression, and other neuropsychiatric disorders is well-documented[9,10] – often, following disasters, resulting in post-traumatic stress disorder (PTSD) and substance use disorders[11]. Moreover, since the same preventive measures meant to limit the spread of an infection can paradoxically also heighten risks due to increased social conflict, isolation, devaluation, rejection, and exclusion, such efforts may be somewhat counterproductive[12]. These effects of increased, sustained psychological stress on individuals who, for various reasons, may be differentially impacted should not be discounted.

Sex is one such factor for concern. Clear sex differences have been shown to exist in exposure to potentially traumatic events and subsequent PTSD[13], and other studies have found females to be more vulnerable to developing mental or physical problems in response to life stressors or potentially traumatic events[14-16]. While early research has suggested that female medical workers may experience or respond more negatively to COVID-19-related events[8], the impact on psychological stress affected has not been fully investigated. This study, then, explores the psychological health and well-being of the general population in China, as well as factors - notably, sex - which may moderate its negative impact.

## Methods

In February of 2020, we administered to the general Chinese population an anonymous, online questionnaire using WeChat, a popular social networking platform. The cross-sectional study utilized a non-random “convenience” or “snowball” sample designed to elicit maximum participation by consenting adults. The study protocol received the appropriate human subjects review and approval.

The survey instrument was designed by several co-authors with psychiatric and epidemiological backgrounds (DL, KS, SY) and, in addition to back socio-demographic information (e.g., sex, age, marital status, living conditions), asked about individuals’ experiences of and reactions to the COVID-19 epidemic – including psychological stress, resilience, and the perceived need for psychological support services. Psychological stress was measured using a 10-centimeter visual analogue scale (VAS) anchored at each end with *“not stressful”* (0) and *“extremely stressful”* (10)[17], The Chinese version of the 10-item Connor-Davidson Resilience Scale (CD-RISC) was used to assess the psychological resilience using items on a 4-point scale ranging from 0 *(“Not at All’)* to 4 *(“Extremely True”)*. For all measures, higher score indicated higher levels of the respective construct[18].

## Statistical Analysis

All analyses were based on 3088 participants, and descriptive statistics included frequency counts and proportions. Multiple linear regression was used to analyze the influence of various independent variables on self-reported psychological stress, including: sex, age (≤ 45, > 45 years), education, occupation, epidemic intensity (provincial prevalence of infections on 03/01/20), existing health conditions (0, 1-2, > 2), local epidemic status(on the rise, at the peak, smoothing, uncertainty), respondent’s infection status (confirmed, suspected, etc.), household size, frequency of personal contact, COVID-19 related health information needs, and mood and emotions experienced. Marital status was found to be highly associated with household size, and was not included in the model. Stepwise model-selection method was used, which the entry and stay significant level of the model were set 0.05.

To explore sex differences, separate regression models were specified for male and female, and independent t-tests were used to assess differences in mean psychological stress and CD-RISC scores. Differences in other measures, such as adaption to current living/working status, coping strategy for healing, and the perceived need for psychological support services, were analyzed using Chi-square tests.

The specified critical p value for all inferential analyses was ≤ 0.05, and all analyses were conducted using Statistical Analysis Software 9.4.3 (SAS institute Inc, Cary, NC).

## Results

A total of 3088 questionnaires were received from residents across 32 Chinese provinces (see Appendix Figure 1). Respectively, females and males comprised 56.5% and 43.5% of respondents; the average age was 37.5 years (SD = 13.5). Almost 1 in 5 (18.5%) were employed as medical personnel – 7.0% of whom reported working on the “front line” of the epidemic. The characteristics of participants were showed in Table 1. The descriptions of each item in the questionnaire were showed in Table 1 of Appendix. The mean self-reported psychological stress score was 3.4 (SD = 2.4). The average psychological resilience score was 28.6 (SD = 8.1) (Table 2).

Results of the multiple regression analysis, using psychological stress as the dependent variable, found several statistically-significant predictors, including being: (1) female; (2) ≤ 45 years old; (3) more highly educated; (4) a farmer/worker/clerical and business/service; (5) unemployed; and (6) in poorer health. Having close contact or completing a medical observation, and a desire for know more about COVID-19 were also positively associated with self-reported stress. Several other independent variables were significantly *negatively* related to psychological stress: (1) frequent contact with colleagues; (2) calmness of mood; and (3) psychological resilience (Table 3).

**Table 1.**
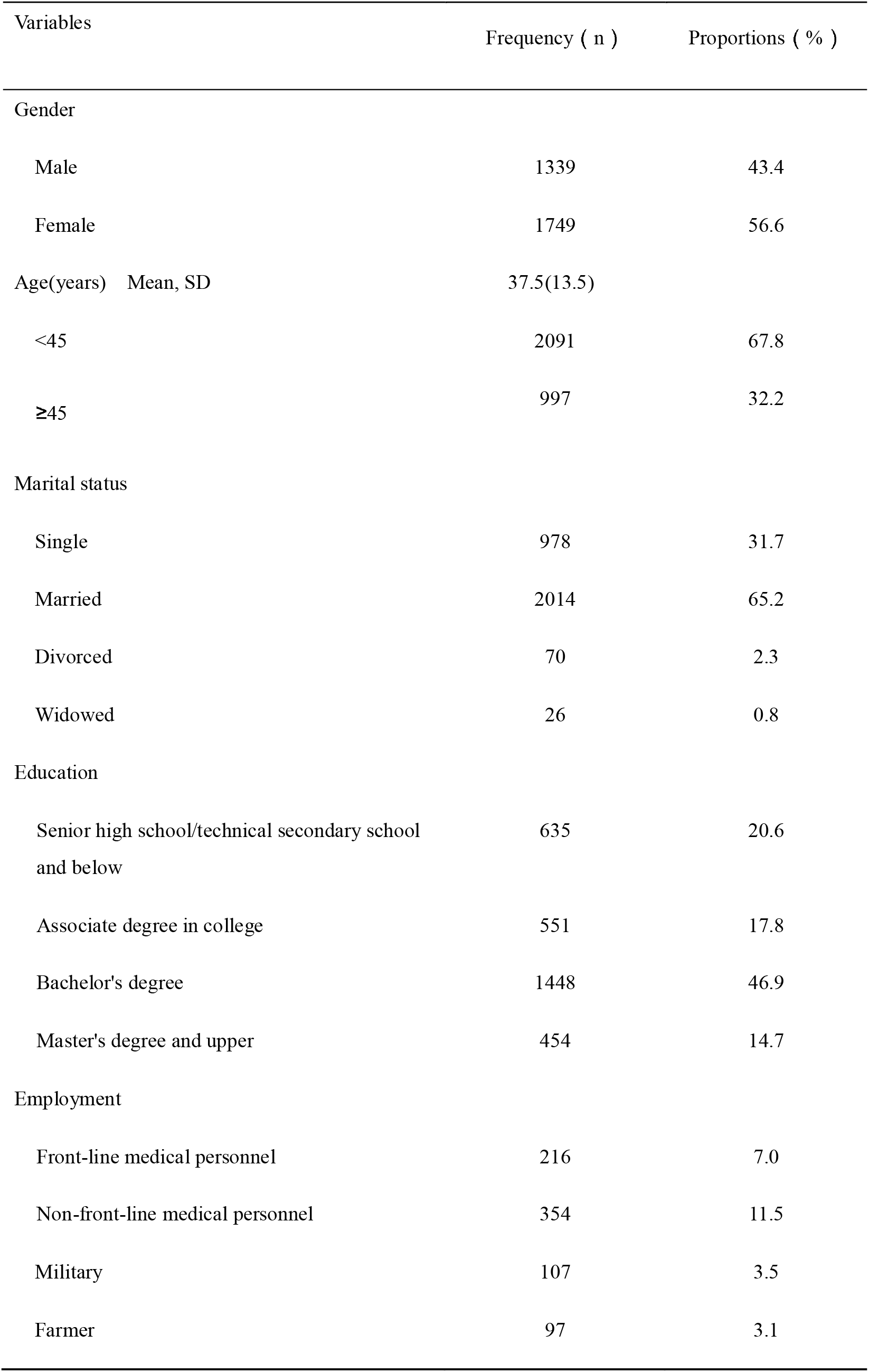
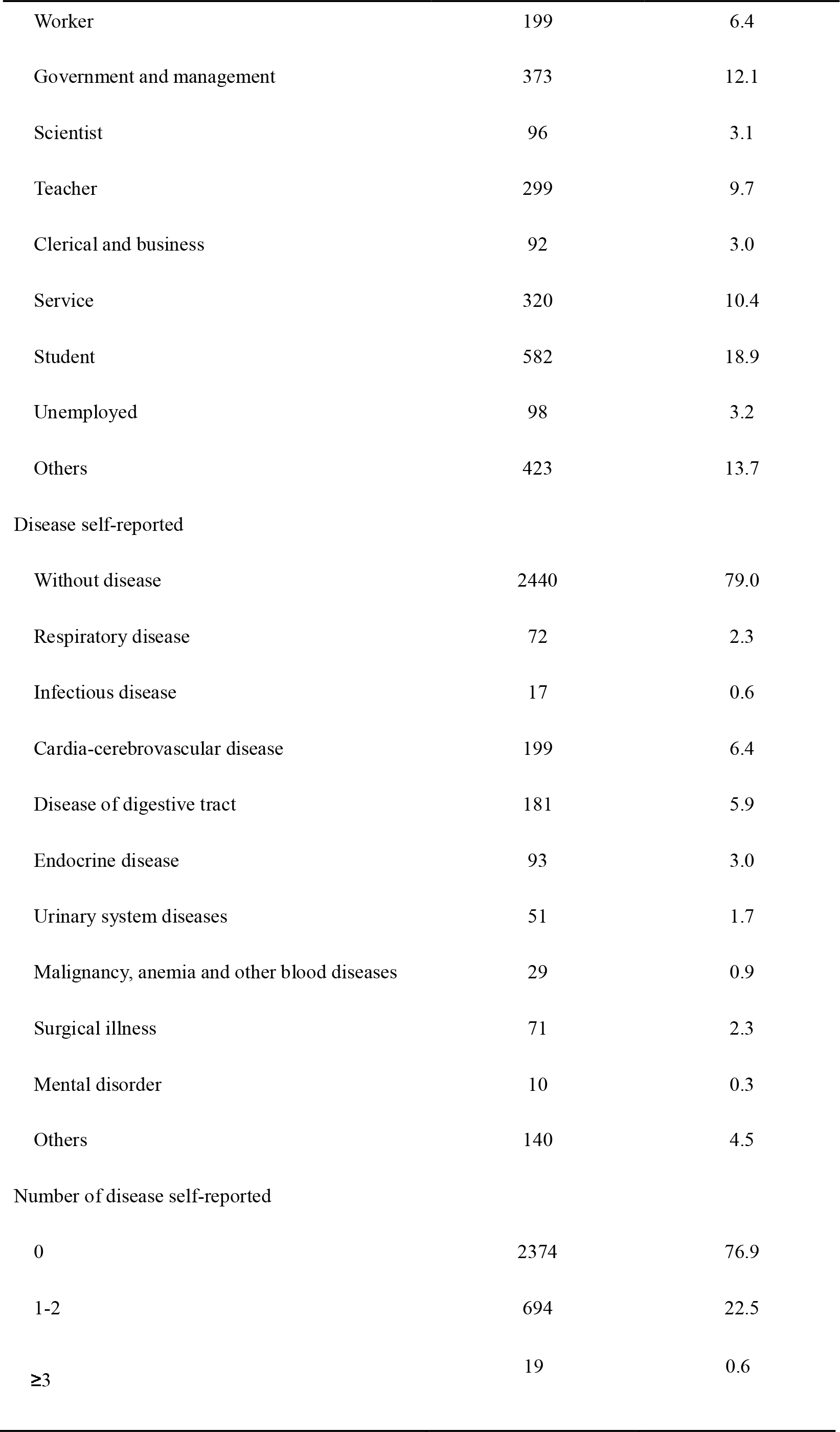
The demographic information of the respondents (n= 3088)

| Variables |  | Frequency ( n ) | Proportions ( % ) |
| --- | --- | --- | --- |
| Gender |  |  |  |
| Male |  | 1339 | 43.4 |
| Female |  | 1749 | 56.6 |
| Age(years) | Mean, SD | 37.5(13.5) |  |
| <45 |  | 2091 | 67.8 |
| ≥45 |  | 997 | 32.2 |
| Marital status |  |  |  |
| Single |  | 978 | 31.7 |
| Married |  | 2014 | 65.2 |
| Divorced |  | 70 | 2.3 |
| Widowed |  | 26 | 0.8 |
| Education |  |  |  |
| Senior high school/technical secondary school and below |  | 635 | 20.6 |
| Associate degree in college |  | 551 | 17.8 |
| Bachelor's degree |  | 1448 | 46.9 |
| Master's degree and upper |  | 454 | 14.7 |
| Employment |  |  |  |
| Front-line medical personnel |  | 216 | 7.0 |
| Non-front-line medical personnel |  | 354 | 11.5 |
| Military |  | 107 | 3.5 |
| Farmer |  | 97 | 3.1 |

|  |  |  |
| --- | --- | --- |
| Worker | 199 | 6.4 |
| Government and management | 373 | 12.1 |
| Scientist | 96 | 3.1 |
| Teacher | 299 | 9.7 |
| Clerical and business | 92 | 3.0 |
| Service | 320 | 10.4 |
| Student | 582 | 18.9 |
| Unemployed | 98 | 3.2 |
| Others | 423 | 13.7 |
| Disease self-reported |  |  |
| Without disease | 2440 | 79.0 |
| Respiratory disease | 72 | 2.3 |
| Infectious disease | 17 | 0.6 |
| Cardia-cerebrovascular disease | 199 | 6.4 |
| Disease of digestive tract | 181 | 5.9 |
| Endocrine disease | 93 | 3.0 |
| Urinary system diseases | 51 | 1.7 |
| Malignancy, anemia and other blood diseases | 29 | 0.9 |
| Surgical illness | 71 | 2.3 |
| Mental disorder | 10 | 0.3 |
| Others | 140 | 4.5 |
| Number of disease self-reported |  |  |
| 0 | 2374 | 76.9 |
| 1-2 | 694 | 22.5 |
| $\geq 3$ | 19 | 0.6 |

**Table 2.**
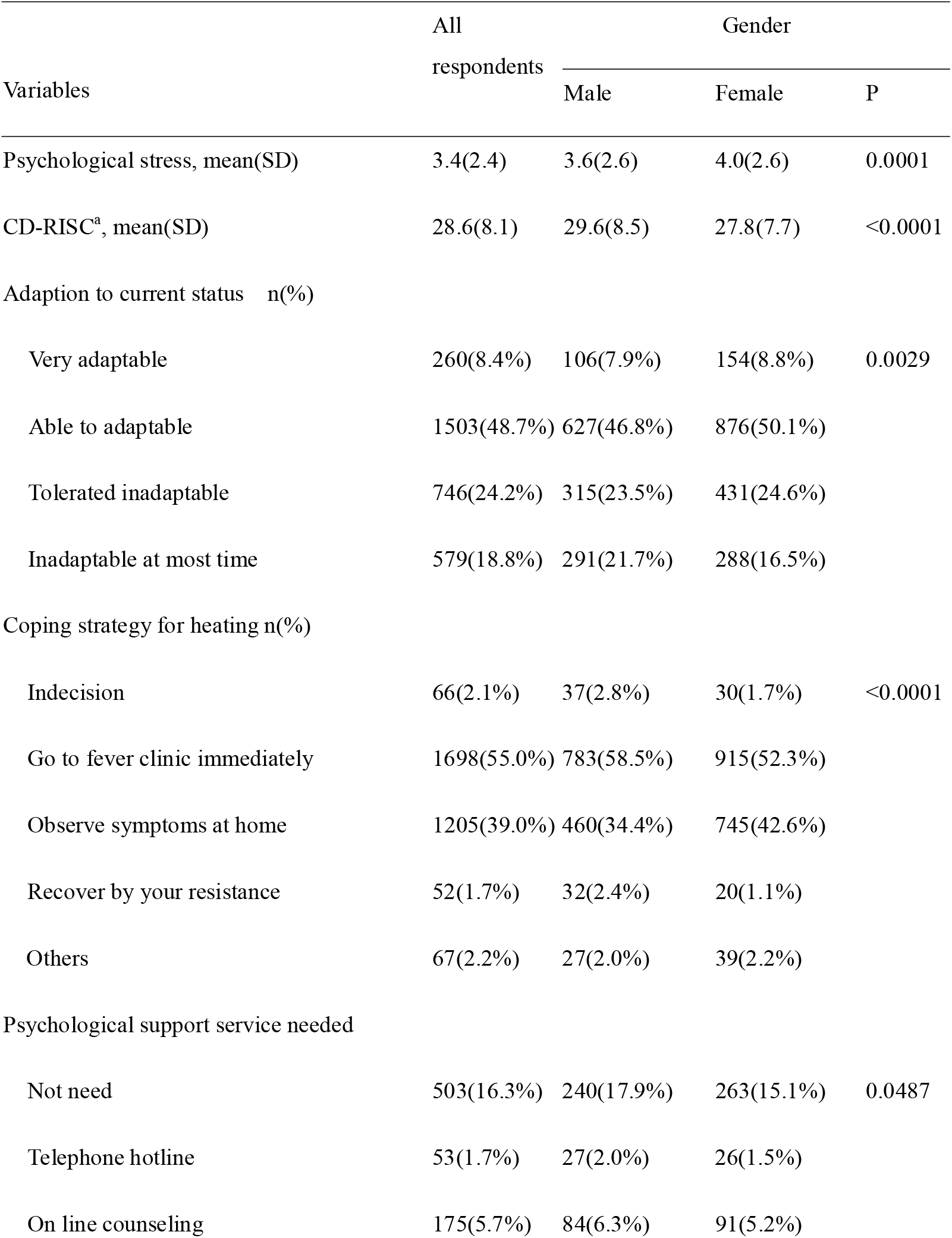
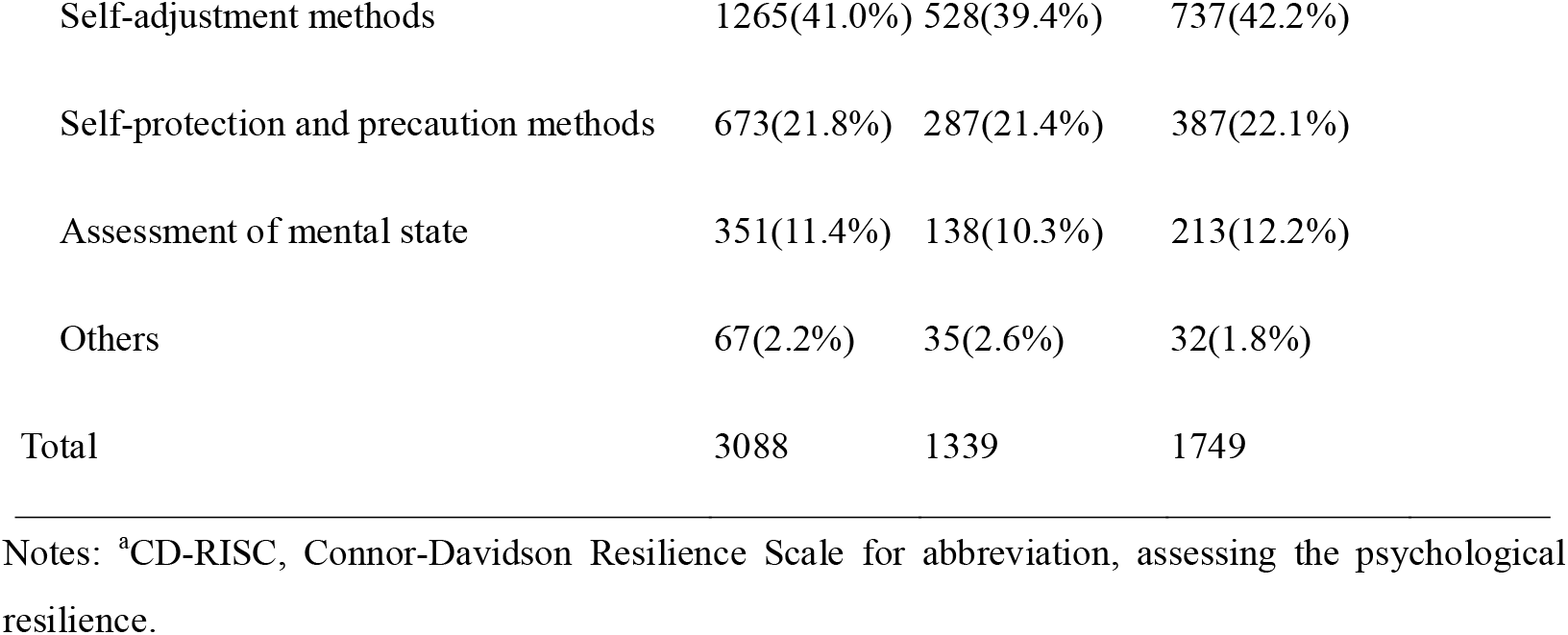
Sex difference on psychology, behaviors and needs to cope with COVID-19

| Variables | All<br>respondents | Gender |  | P |
| --- | --- | --- | --- | --- |
|  |  | Male | Female |  |
| Psychological stress, mean(SD) | 3.4(2.4) | 3.6(2.6) | 4.0(2.6) | 0.0001 |
| CD-RISC<sup>a</sup>, mean(SD) | 28.6(8.1) | 29.6(8.5) | 27.8(7.7) | <0.0001 |
| Adaption to current status n(%) |  |  |  |  |
| Very adaptable | 260(8.4%) | 106(7.9%) | 154(8.8%) | 0.0029 |
| Able to adaptable | 1503(48.7%) | 627(46.8%) | 876(50.1%) |  |
| Tolerated inadapttable | 746(24.2%) | 315(23.5%) | 431(24.6%) |  |
| Inadapttable at most time | 579(18.8%) | 291(21.7%) | 288(16.5%) |  |
| Coping strategy for heating n(%) |  |  |  |  |
| Indecision | 66(2.1%) | 37(2.8%) | 30(1.7%) | <0.0001 |
| Go to fever clinic immediately | 1698(55.0%) | 783(58.5%) | 915(52.3%) |  |
| Observe symptoms at home | 1205(39.0%) | 460(34.4%) | 745(42.6%) |  |
| Recover by your resistance | 52(1.7%) | 32(2.4%) | 20(1.1%) |  |
| Others | 67(2.2%) | 27(2.0%) | 39(2.2%) |  |
| Psychological support service needed |  |  |  |  |
| Not need | 503(16.3%) | 240(17.9%) | 263(15.1%) | 0.0487 |
| Telephone hotline | 53(1.7%) | 27(2.0%) | 26(1.5%) |  |
| On line counseling | 175(5.7%) | 84(6.3%) | 91(5.2%) |  |

|  |  |  |  |
| --- | --- | --- | --- |
| Self-adjustment methods | 1265(41.0%) | 528(39.4%) | 737(42.2%) |
| Self-protection and precaution methods | 673(21.8%) | 287(21.4%) | 387(22.1%) |
| Assessment of mental state | 351(11.4%) | 138(10.3%) | 213(12.2%) |
| Others | 67(2.2%) | 35(2.6%) | 32(1.8%) |
| Total | 3088 | 1339 | 1749 |

Notes: <sup>a</sup>CD-RISC, Connor-Davidson Resilience Scale for abbreviation, assessing the psychological resilience.

**Table 3.** Multiple linear regression for psychological stress and its influence factors

| Variables | All respondents |  | Male |  | Female |  |
| --- | --- | --- | --- | --- | --- | --- |
|  | Estimates | P values | Estimates | P values | Estimates | P values |
| Intercept | 7.4 | <.0001 | 6.9 | <.0001 | 9.4 | <.0001 |
| sex | 0.3 | 0.0036 | - | - | - | - |
| Age(<45, ≥45 years) | -0.4 | <.0001 | -0.4 | 0.0078 | -0.4 | 0.0009 |
| Education | 0.2 | <.0001 | 0.3 | <.0001 | - | - |
| Employment(Military as reference) |  |  |  |  |  |  |
| Unsteady income<sup>a</sup> | 0.4 | 0.0002 | 0.4 | 0.007 | - | - |
| Unemployed | 0.6 | 0.0211 | - | - | - | - |
| Number of diseases self-reported | 0.3 | 0.0041 | - | - | 0.4 | 0.0077 |
| Local epidemic status | - | - | - | - | -0.3 | 0.0059 |
| Risk level of the population <sup>b</sup> | -0.9 | 0.0038 | -1.2 | 0.0057 | - | - |
| Contacted more than 3 times a week with colleagues | 0.3 | 0.0013 | - | - | 0.4 | 0.0007 |
| Desire to acquire knowledge of COVID-19 | -0.2 | 0.0006 | - | - | -0.3 | <.0001 |
| Time spent on the outbreak in one day | 0.1 | 0.0396 | 0.2 | 0.007 | - | - |
| Mood changes comparing with the early epidemic phase | -0.4 | <.0001 | -0.4 | <.0001 | -0.4 | <.0001 |
| Difficulties encountered (Social limitations as reference) |  |  |  |  |  |  |
| Problem of diseases | 0.5 | 0.0235 | - | - | 0.8 | 0.0086 |
| Psychological problems | 1.2 | <.0001 | 1.1 | 0.0002 | 1.3 | <.0001 |
| Economic problems | 0.8 | <.0001 | 0.8 | 0.0004 | 0.9 | <.0001 |
| Unable to work/study | 0.3 | 0.0072 | - | - | 0.3 | 0.0357 |
| CD-RISC | -0.1 | <.0001 | -0.1 | <.0001 | -0.1 | <.0001 |

Notes: <sup>a</sup> Unsteady income were including farmer, worker, clerical, business and service; <sup>b</sup> Close contact or completed a medical observation vs. general people

## Male and Female Stressors

Except for age, high education, and resilience, for male, unstable income population (farmer/worker/clerical and business/service), more time spent on the outbreak, risk infection (close contact or completed a medical observation), psychological and economic problems aggravated psychological stress. In female, poorer health, worse local epidemic status, higher desire for knowledge about the COVID-19, problem of diseases during the epidemic and unable to work/study were the risk factors. Calmness mood and frequently contacting with colleagues were the protective factor of stress.(Table 3).

Sex differences did exist with regard to adaptation, responses to potential symptoms, and the perceived need for psychological support services – with males being less adaptive and less likely to see a need for psychological support, but more likely to seek immediate medical attention for suspected feverish symptoms (see Table 2).

## Discussion

Research as consistently shown that major life changes can lead to severe and sometimes chronic psychological stress[17]. With more than 2.1 cases confirmed worldwide, and over 140,000 reported deaths[19], the COVID-19 epidemic constitutes a pervasive source of potential stress on a global scale. Indeed, with many countries swiftly instituting strict control measures, normal routines were drastically disrupted with the closing of businesses, industries, and schools – and the recommendation (or requirement) that individuals remain at home. Such behavioral changes, whether mandatory or not, can be expected to negatively challenge individuals’ mental health and/or emotional well-being.

Our study, conducted across provincial China (Taiwan excepted) at the height of the COVID-19 epidemic, suggests that being female, somewhat younger (≤ 45 years old), more highly educated, unemployed, and in poorer overall health were all risk factors for experiencing psychological stress. Uncertainty of one’s local disease status (epidemic) and some prior personal COVID-19-related contact were also factors contributing to one’s perceived stress. In contrast, “protective” factors included frequent contacting with colleagues, calmness of mood, and psychological resilience. Higher desire for knowledge about the COVID-19, diseases/psychological/economic problems and cannot go to work/study difficulties during the epidemic were the risk factors of stress.

Disease susceptibility, and the economic problems that can result from an inability to work, can be prime contributors to psychological stress. Similarly, uncertainty and lack of control resulting from lockdowns, restrictions, quarantines, etc. arguably impacted every Chinese residents’ life and, potentially, detrimentally affected their physical, social-psychological, and economic conditions[2,3], The sustained, long-term implementation of these safeguards undoubtedly prolonged an already stressful and challenging situation – and the loss of both income and personal identity associated with the lack of employment likely resulted in increased anxiety. Indeed, for respondents without sustainable incomes, the effects were especially dire.

The full range of possible impacts should be considered when implementing disease control and prevention measures, and disseminating easily-assessible, understandable knowledge of COVID-19 and providing alternative venues for personal contact with friends or colleagues can be effective buffers against psychological stress. Conforming to the common perceptions, people alerted to risks and threats instinctively seek outside help, confirmed in a recent Chinese study demonstrating that individuals, on average, spent ≥ 3 hours per day during the epidemic associated with mental health [20], Social support, typically associated with lower depression and anxiety, could further buffer the cognitive effects of stress[21]. Our findings suggest that appropriate social supports, such as frequent contacting with colleagues, to relieve stress during an epidemic might include providing more professional knowledge of protective measures, real-time updates and report, access to urgent medical service, basic living security measures, and alternative means to interpersonal communication.

Age was another factor related to self-reported stress – with study findings suggesting that younger (≤ 45) respondents experienced greater stress. These results were consistent with the previously-cited Chinese COVID-19 study and others involving the psychological impacts of disasters[20,22]. Exactly why this is the case remains somewhat unclear: Perhaps older persons pay more attention on positive emotion simulation and neglect negative simulation (“positive effects”)[23], or maybe those younger face feel greater social, emotional, and/or economic responsibilities toward their families’ health and protection.

Our study also highlighted the importance of resilience as a “protective” factor to psychological stress, often vis-à-vis a greater sense of adaptability and control over one’s external environment. In fact, studies have found that psychological resilience both directly and indirectly protects some individuals against stress-related mental health problems (e.g., PTSD, anxiety, depression)[24,25]. In our study, males’ higher resilience may partially explain their comparatively (vs. females) lower stress levels.

This finding of female’s stress was being greater than that of males in response to such situations was also consistent with existing evidence[14,16]. Observed sex differences of stress are often attributed to differential impacts on individuals’ social environmental, psychodynamic, and cognitive processes[26,27]. Behavioral responses to distress and the experience/expression of emotion are also thought to be moderated by sex[13] and, more recently, sex differences in susceptibility to stress have been expanded to include physiological factors [28,29] such as ovarian hormone fluctuations[26,30] and endogenous estradiol changes across the menstrual cycle [31], Similarly, stress-related fMRI studies have found brain functions associated with emotion and stress regulation, self-referential processing, and cognitive control to be more pronounced in males[32].

Sex differences in self-reported stress are further reflected in the perceived need of psychological support services, which were seen as more evident in females than in males. Again, the underlying mechanisms are unclear, but it may be that males are more likely to self-manage coping responses to stress, while female are more likely to seek external or professional help.

This study had several limitations. First, although study respondents reflect a national sample, most of the non-random, convenience sample were located outside the heaviest epidemic area – which may have had some impact on findings. Moreover, the cross-sectional design makes establishing the causal nature of relationships problematic. Second, in response to the rising trend of COVID-19 epidemic and the infectious characteristics, we were forced to use WeChat online survey. Finally, for ethical reasons, we purposely did not ask about confirmed or suspected infection among respondents themselves, and the proportion reporting close contact and having completed medical observations were few.

## Conclusion

Self-reported stress, as measured, was significantly related to sex, age, education, and employment during the epidemic of COVID-19. Findings suggest responses to future emergency situations (e.g., disease epidemics) should bolster appropriate control measures with targeted means of psychological support.

## Data Availability

All data are available from the corresponding author.

## Fundings

This study was funded by the National Natural Science Foundation of China (81973713), State Administration of Traditional Chinese Medicine of China Support Project (2017ZX10106001), Logistics Research Project (ALB18J002, BHJ15J003), and Chinese Medical Scientific Development Foundation of Beijing City (JJ2018-101), Project of Institute of Basic Research in Clinical Medicine, China Academy of Chinese Medical Sciences (Z0652).

## Conflict of Interest

There was no conflict of interest information.

## Availability of data and material

The data and material can be accessed from

## Code availability

The code can be accessed from.

## Authors’ contribution

Shiyan Yan, Kangxin Song, Dan Luo, and Fengsu Hou design the study. Kangxin Song, Dan Luo, and Fengsu Hou wrote the study protocol. Qizhen Wang, Fengsu Hou, FengyingBi, Dan Luo, Rong Jiao and Kangxin Song collected the data. Shiyan Yan carried out the analysis with support from Terry D. Stratton, Voyko Kavcic and Yang Jiang. Rui Xu and Shiyan Yan wrote the draft of the manuscript. Terry D. Stratton, Voyko Kavcic and Yang Jiang revised the draft. All authors contributed to the final version.

## Ethics approval

The study has been proved by the institution review board of School of Public Health, Central South University(XYGW-2020-04).

## Notes

### Competing Interest Statement

The authors have declared no competing interest.

### Author Declarations

XYGW-2020-04

